# Causal effects of motor control on gait kinematics after orthopedic surgery in cerebral palsy: a machine-learning approach

**DOI:** 10.1101/2022.01.04.21268561

**Authors:** Katherine M. Steele, Michael H. Schwartz

## Abstract

**Background:** Altered motor control is common in cerebral palsy (CP). Understanding how altered motor control effects movement and treatment outcomes is important, but challenging due to complex interactions between impairments. While regression can be used to examine associations between impairments and gait, causal modeling provides a mathematical framework to specify assumed causal relationships, identify covariates that may introduce bias, and test model plausibility. The goal of this research was to quantify the causal effects of altered motor control and other impairments on gait, before and after single-event multi-level orthopedic surgery (SEMLS).

**Methods:** We evaluated the impact of SEMLS on change in Gait Deviation Index (ΔGDI) between gait analyses. We constructed our causal model with a Directed Acyclic Graph that included the assumed causal relationships between SEMLS, ΔGDI, baseline GDI (GDI_pre_), baseline neurologic and orthopedic impairments (Imp_pre_), age, and surgical history. We identified the adjustment set to evaluate the causal effect of SEMLS on ΔGDI and the impact of Imp_pre_ on ΔGDI and GDI_pre_. We used Bayesian Additive Regression Trees (BART) and accumulated local effects to assess relative effects.

**Results:** We prospectively recruited a cohort of children with bilateral CP undergoing SEMLS (N=54, 35 males, age: 10.5±3.1 years) and identified a control cohort with bilateral CP who did not undergo SEMLS (N=55, 30 males, age: 10.0±3.4 years). There was a small positive causal effect of SEMLS on ΔGDI (1.68 GDI points). Altered motor control (*i*.*e*., dynamic and static motor control) and strength had strong effects on GDI_pre_, but minimal effects on ΔGDI. Spasticity and orthopedic impairments had minimal effects on GDI_pre_ or ΔGDI.

**Conclusions:** Altered motor control and other baseline impairments did have a strong effect on GDI_pre_, indicating that these impairments do have a causal effect on a child’s gait pattern but minimal effect on expected changes in GDI after SEMLS. Heterogeneity in outcomes suggests there are other factors contributing to changes in gait. Identifying these factors and employing causal methods to examine the complex relationships between impairments and movement will be required to advance our understanding and care of children with CP.

## 1 Introduction

Children diagnosed with cerebral palsy (CP) exhibit altered motor control due to an injury to the brain at or near the time of birth^1-3^. Altered motor control can be observed in CP in many ways, such as increased co-contraction, decreased capacity to selectively move individual joints, decreased complexity of control, and altered movement patterns. Prior research has suggested that quantifying motor control is important to understand function and inform treatment planning ^4-8^. However, altered motor control occurs and interacts with many other impairments in CP, which makes quantifying and isolating the effects of altered motor control challenging.

In addition to altered motor control, children with CP often have other neurologic impairments such as spasticity, dystonia, or muscle weakness. Over time, orthopedic impairments can also develop, including muscle contractures and altered bone morphology^9-13^. Together, these neurologic and orthopedic impairments contribute to limitations in movement, which reduce walking speed, elevate walking energy cost, and limit the capacity of children with CP to participate in daily activities^14-18^.

The complexity of CP makes it challenging to objectively determine the causal effects of specific impairments, such as altered motor control, on gait. As a result, many children with CP undergo clinical gait analysis (CGA)^19^, which provides quantitative measures of a child’s gait pattern that can be tracked over time and used to inform treatment decisions^20-22^. In particular, CGA was historically developed to support decision making for orthopedic surgery^23-26^. Many children’s hospitals now have CGA laboratories used for pre-operative and post-operative assessments.

While CGA has been used for treatment planning for over 30 years, deciphering causal effects of impairments on gait has remained elusive. Data from CGA is traditionally used to evaluate associations between a specific impairment and an outcome measure, typically using bivariate or multivariate regression analyses applied to retrospective data^27-31^. In cases where multivariate regression has been used, the choice of variables for inclusion has often not had a clear causal basis. Our prior work to evaluate the impact of motor control on gait and treatment outcomes have relied on these methods^5,6,32^. Using multivariate regression with retrospective data from multiple hospitals we have repeatedly demonstrated that Dynamic Motor Control (DMC) during walking is associated with function (*i*.*e*., Gross Motor Functional Classification System Levels or Gillette Functional Assessment Questionnaire) and outcomes (*i*.*e*., Gait Deviation Index, Walking Speed, Pediatric Outcomes Data Collection Instrument) after orthopedic surgery, rhizotomy, or botulinum toxin injections^6,32^. Similar analyses have demonstrated that other impairments – such as strength, hamstring length, motor control, or torsional deformities – are also correlated with treatment outcomes^33-37^.

Understanding whether altered motor control or other impairments *cause* altered gait or treatment outcomes is nearly impossible with non-causal regression alone. Given the complexity and heterogeneity of CP, this “*implied cause by association*” approach, without regard to possible confounding, is likely to lead to confusing and even erroneous conclusions. For example, researchers may observe that strength is associated with walking speed. However, strength is also affected by other primary neurologic deficits, like poor motor control, which may have an independent causal impact on speed. Understanding causal effects is impossible without considering these causal pathways and adjusting for relevant factors.

In recent years, there has remarkable growth in the development and successful applications of causal inference methods^38,39^. From a conceptual perspective, causal methods allow researchers to explicitly share assumed causal relationships and mathematically define covariates necessary for estimating causal effects^40^. From a computational perspective, numerous algorithms have been developed for modeling causal outcomes. Among the most successful of these are Bayesian Additive Regression Trees (BART), which have been shown to produce estimates of causal effects with low levels of bias and variance, and realistic confidence intervals^41-44^. Williams and colleagues (2018) have highlighted the potential of causal inference for pediatrics^45^. However, these methods have had limited application in CP or biomechanics research.

The goal of this research was to quantify the causal effects of motor control and other impairments on gait, before and after orthopedic surgery. Specifically, we prospectively recruited children with CP who were undergoing single-event multilevel orthopedic surgery (SEMLS). We also identified a cohort of controls from the same time period who were not undergoing SEMLS between gait analyses. We developed a causal model and used BART to quantify the effects of motor control and other impairments on changes in gait kinematics after SEMLS. These methods provide a foundation for understanding the complex and interactive effects of impairments on gait for children with CP.

## 2 Methods

### 2.1 Participants

We recruited children with bilateral CP who were between 6 and 18 years old at the time of baseline gait analyses and scheduled for SEMLS. The goal of our prospective recruitment was to follow a representative cohort of patients at Gillette Children’s Specialty Healthcare from their baseline gait analysis through two follow-up assessments at six-months and one-year after SEMLS. The one-year analysis was our primary outcome; however, the COVID-19 pandemic interrupted the one-year follow-up for two participants, and we used the six-month follow-up visit for these participants. We included patients whose baseline gait analysis was no more than six months before their scheduled surgery date. We defined SEMLS as surgery consisting of two or more major orthopedic procedures on a single side. We also identified a cohort of controls with CP who did not undergo SEMLS. We identified children with bilateral CP who underwent multiple gait analyses with kinematic and electromyographic (EMG) recordings, with a maximum time of 2.5 years between visits during the same time period. We excluded participants who underwent prior or current rectus femoris transfer, since we were evaluating motor control from EMG recordings. This research was conducted with approval from the University of Minnesota Institutional Review Board.

### 2.2 Causal Model

For this analysis we focused on evaluating the impact of SEMLS on gait kinematics. We *a priori* specified our outcome measure as the Gait Deviation Index (GDI, ClinicalTrials.gov NCT02699554) as a common summary measure of walking kinematics that has been used extensively in prior studies to evaluate and predict treatment outcomes.

We constructed our causal model with a Directed Acyclic Graph (DAG)^46-48^. The logic behind our DAG is as follows (Figure 1):

**Figure 1:**
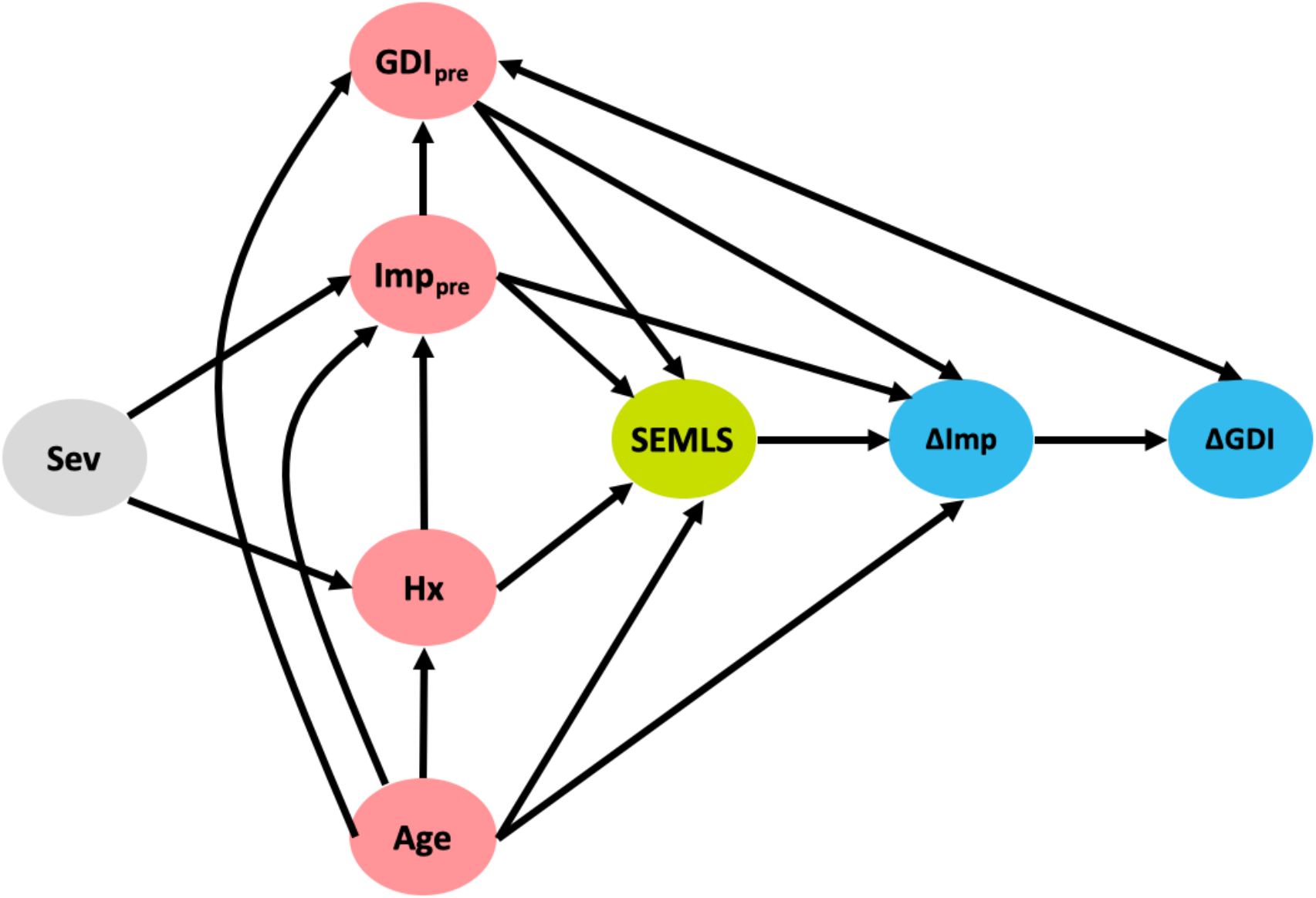
DAG describing the assumed causal relationships between SEMLS (exposure) and ΔGDI (outcome). The causal relationship between SEMLS and ΔGDI is mediated by changes in impairments (ΔImp). Baseline GDI (GDI_pre_), baseline impairments (Imp_pre_), surgical history (Hx), and Age are also included as causal factors. The DAG also includes unmeasured factors related to general CP severity, which impact baseline impairments and surgical history. The step-by-step process and rationale for this DAG are available in the Supplementary Material (http://dagitty.net/mUCSPWo).

1. Our objective was to determine the impact of SEMLS on change in GDI (ΔGDI). Thus, SEMLS is our exposure and ΔGDI is our outcome. SEMLS induces a change in impairments (ΔImp) that causes the observed ΔGDI.
2. The covariates we identified as common causes of both SEMLS (*i.e*., variables that impact the choice to undergo SEMLS) and ΔGDI included: Age, baseline GDI (GDI_pre_), and baseline impairment (Imp_pre_). Baseline impairments represent a set of variables collected during CGA to evaluate neurologic and orthopedic impairments (Table 1).

**Table 1:** Variable definitions
3. Surgical treatment history (Hx) is a common cause of baseline impairment (Imp_pre_) and whether or not SEMLS is recommended.
4. We included a general severity (Sev) measure as an unmeasured factor that impacts baseline impairment (Imp_pre_) and surgical treatment history (Hx).

| Variable | Description |
| --- | --- |
| GDI | Overall measure of the deviation in an individual's kinematics compared to nondisabled peers scaled such that mean(sd) over the nondisabled population is 100(10)<sup>51</sup>. Kinematics were evaluated using marker-based motion analysis and a modified plug-in-gait marker set. |
| SEMLS | Binary variable indicating whether or not child had single-event multi-level orthopedic surgery, defined as a surgery with two or more orthopedic surgeries on at least one leg. |
| Hx | Binary list of prior surgical treatments. |
| Age | Years from birth defined as days/365.25 |
| Impairments | **Spasticity:** Mean modified Ashworth score across plantarflexors, hamstrings, hip adductors, and rectus femoris. |
|  | **Strength:** Mean manual muscle strength score across hip flexors/extensors, knee flexors/extensors, and ankle dorsiflexors/plantarflexors where 1 is defined as a 'visible or palpable contraction' and 5 is defined as 'full range of motion against gravity'. |
|  | **Static Motor Control (SMC):** Mean static motor control score across hip abduction, hip flexion, hip extension, knee extension, and ankle plantarflexion where 0 is very little or no control of single joint movement, 1 is impaired voluntary movement at a single joint, and 2 is good voluntary movement at a joint. |
|  | **Dynamic Motor Control (DMC):** Measure of the complexity of muscle activity during gait evaluated from synergy analysis of EMG data. Complexity is evaluated as the total variance accounted for by one synergy of EMG data during CGA and compared to nondisabled peers scaled such that mean(sd) over the nondisabled population is 100(10)<sup>5,52</sup>. |
|  | **Torsional Deformity:** Femoral anteversion and tibial torsion (bimalleolar axis angle) measured during physical exam. |
|  | **Contracture:** Measures of joint range of motion from physical exam including maximum ankle dorsiflexion with the knee extended, maximum knee extension, unilateral popliteal angle, and maximum hip extension measured during the Thomas Test. |

Note that similar DAGs could be constructed for other outcome measures such as walking speed or energy cost. Similarly, additional factors could be added to the DAG, if there were rational arguments that they were common causes of one of the variables in the DAG and ΔGDI. The step-by-step process we used to construct our DAG is illustrated in the Supplementary Material.

From the DAG we determined the variables that needed to be included in any model (*e.g*., regression, BART) to evaluate the total causal effect of SEMLS on ΔGDI. These variables are called the adjustment set, representing the confounding covariates that could produce bias if not included in an analysis. For this DAG, the minimal sufficient adjustment set to estimate the total causal effect of SEMLS on ΔGDI was: Age, GDI_pre_, and Imp_pre_. We also determined the adjustment set to evaluate the total causal effect of baseline impairment (Imp_pre_) on ΔGDI and GDI_pre_. The minimal sufficient adjustments sets were Age and Hx for ΔGDI and Age for GDI_pre_. The plausibility of a DAG can be evaluated by identifying conditional independencies, variables that should be independent given the causal relationships defined in the DAG. We identified the adjustment sets and independencies with dagitty^49^ and all analyses were conducted in R (version 4.1.0)^50^.

### 2.3 Bayesian Additive Regression Trees

To assess the total causal effects of SEMLS and baseline impairments (Imp_pre_) on change in GDI (ΔGDI) we used Bayesian Additive Regression Trees (BART), a machine learning method that uses a boosted ensemble of regression trees for nonparametric function estimation relying on a Bayesian probability model^41^. Like other tree-based regression methods, an advantage of BART is that it can handle nonlinear effects and interactions^53^. For causal modeling, recent work has demonstrated that BART-based models achieve accurate and precise causal predictions^43,44^.

For this analysis, we used BART models to estimate ΔGDI using the adjustment sets identified by the DAG. Thus, to identify the impact of SEMLS on ΔGDI, we included the covariates Age, GDI_pre_, and Imp_pre_. Baseline impairments were not available for all participants. Missing data in Imp_pre_ were imputed using multivariate imputation by chained equations (MICE)^54^. We used the *bartMachine* package to implement the analysis^55^. We optimized the hyperparameters for each BART model using 10-fold cross-validation. We report the pseudo-R^2^ (*1 – SSE/SST)* for each BART model and used k-fold cross-validation (*k* = 10) to determine the out-of-sample root mean square error (RMSE).

To assess the relative effects of individual variables from BART, we used accumulated local effect (ALE) analysis^56^. The ALE analysis is similar to a partial dependence plot, but the averaging is done locally to avoid including observations that are unlikely to ever be realized (*e.g*., someone walking three standard deviations slower than average but with a normal cadence). The ALE plots illustrate the impact of each variable over the range of values for that variable, conditioned on the other covariates in the model. Thus, ALE plots can be useful for examining nonlinear effects identified by BART. For example, the ALE plot can highlight nonlinear effects such as when a variable impacts GDI with a deviation from average (*i.e*., a U-shaped plot) or when a variable only impacts GDI above or below a certain cut-off (*i.e*., a step function or discontinuity).

## 3 Results

### 3.1 Participants

We prospectively recruited 54 children with bilateral CP who underwent SEMLS (Table 2). During this same time period, we identified 55 children with bilateral CP who visited the gait laboratory for repeat visits and no intervening surgical procedures. The participants who underwent SEMLS were older and had more femoral anteversion, more tibial torsion, and lower GDI scores at the initial gait analysis than the patients who did not undergo SEMLS. The SEMLS participants received, on average, five procedures (Figure 2).

**Table 2:** Participant characteristics, average (SD)

|  | No SEMLS | SEMLS |
| --- | --- | --- |
| **N** | 55 | 54 |
| **Males N** | 30 | 35 |
| **Age (years)** | 10.0 (3.4) | 10.5 (3.1) |
| **GDI** | 69.4 (10.0) | 68.8 (12.1) |
| **GDI Post** | 69.2 (11.9) | 71.4 (11.7) |
| **SMC** | 1.23 (0.42) | 1.11 (0.40) |
| **DMC** | 81.1 (9.0) | 80.6 (9.6) |
| **Strength** | 3.36 (0.59) | 3.55 (0.67) |
| **Spasticity** | 1.16 (0.42) | 1.30 (0.46) |
| **Anteversion (°)** | 36.3 (10.4) | 39.6 (11.3) |
| **Bimalleolar (°)** | 12.6 (10.6) | 13.4 (11.4) |
| **Dorsiflexion (°)** | 1.16 (7.90) | -0.52 (7.48) |
| **Knee Extension (°)** | 0.52 (6.60) | 0.22 (7.51) |
| **Thomas Test (°)** | 0.61 (6.23) | 2.25 (6.16) |
| **Popliteal Angle (°)** | 51.5 (15.4) | 55.9 (12.9) |

**Figure 2:**
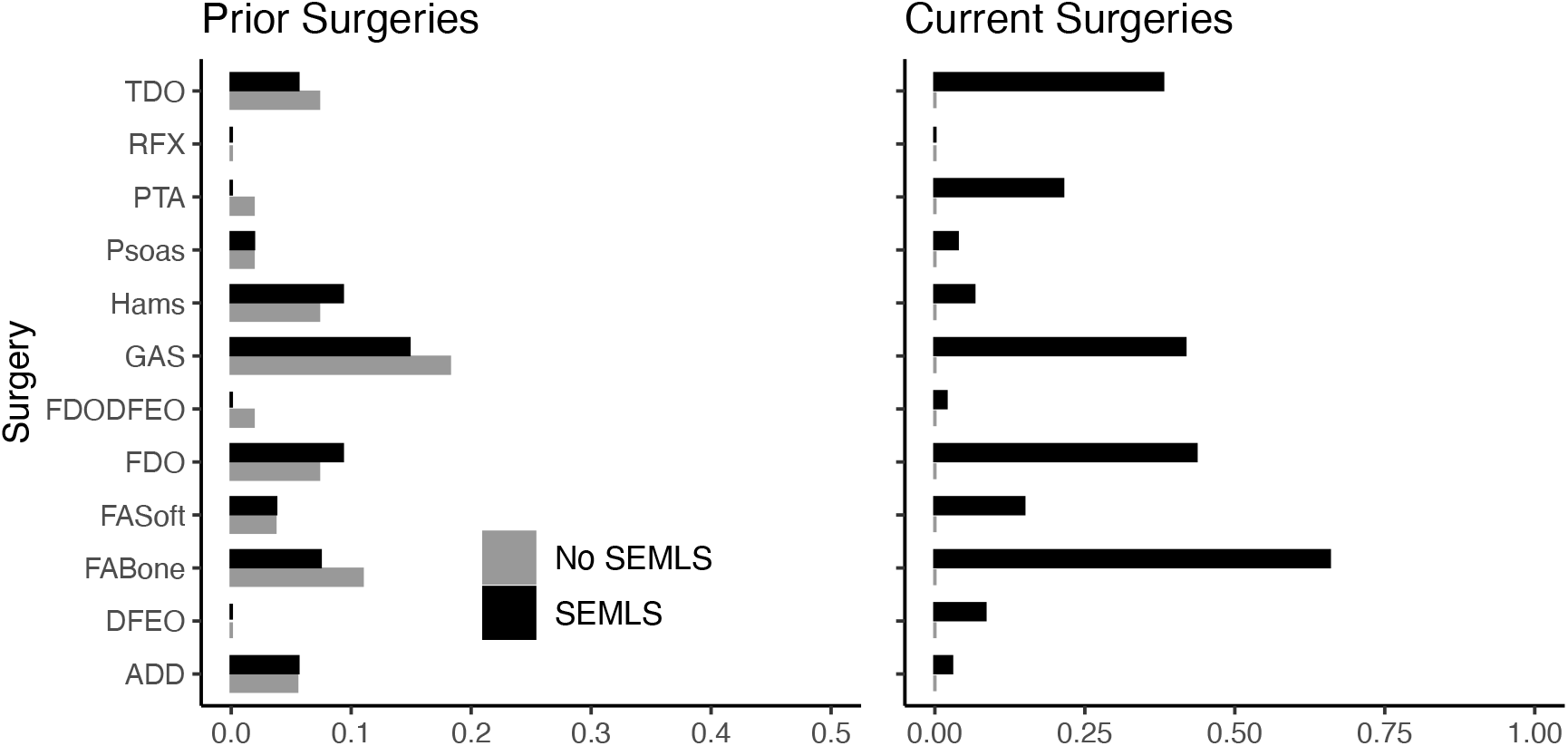
Prior and current surgeries of participants in both cohorts. Note that we excluded potential participants who underwent rectus femors transfer. TDO = tibial derotation osteotomy, RFX = rectus femoris transfer, PTA = patellar tendon advancement, Psoas = psoas lengthening or release, Hams = hamstring lengthening, GAS = plantarflexor lengthening, FDODFEO = distal femoral derotation and extension osteotomy, FDO = femoral derotation osteotomy, FAsoft = foot/ankle soft tissue procedure, FABone = foot/ankle boney procedure, DFEO = distal femoral extension osteotomy, ADD = adductor lengthening or release.

### 3.2 Effects of SEMLS

There was a small positive causal effect of SEMLS on ΔGDI. The estimated total causal effect of SEMLS on ΔGDI was 1.68 GDI points, representing the difference between the SEMLS (+0.85 GDI points) and control (−0.83 GDI points) cohorts. While the average change in GDI between visits was 2.64±8.12 for the SEMLS cohort and -0.26±7.44 for the control cohort, the total causal effects represents the estimated effect of SEMLS after adjusting for differences in Age, GDI_pre_, and Imp_pre_. The BART model explained 18% of the variance in ΔGDI, with an out-of-sample root mean square error of 7.72. The implied conditional independencies of the DAG were also evaluated and all partial correlations were less than 0.20, supporting model plausibility (Supplementary Material).

### 3.3 Effects of Impairments

Baseline values of neurologic and orthopedic impairments (Imp_pre)_ had minimal effects on ΔGDI (Figure 3). SMC, DMC and strength had moderate effects on GDI_pre_, but not ΔGDI. Greater SMC or DMC resulted in higher GDI_pre_ scores, while muscle weakness had a negative impact on GDI_pre_ scores. Orthopedic impairments had smaller effects on GDI_pre_. Knee extension range of motion and tibial torsion (*i.e*., bimalleolar angle) had the largest effect among orthopedic impairments on GDI. Participants who had excessive knee range of motion (*i.e*., hyperextension) had worse baseline GDI scores. Contracture of the plantarflexors, hamstrings, or iliopsoas, as well as femoral anteversion had minimal impact on GDI_pre_ or ΔGDI. The BART models evaluating the effects of impairments explained 61% of the variance in GDI_pre_ and 9% of the variance in ΔGDI. Out-of-sample performance of the BART models were able to RMSE = 9.02 for GDI_pre_ and RMSE = 7.97 for ΔGDI.

**Figure 3:**
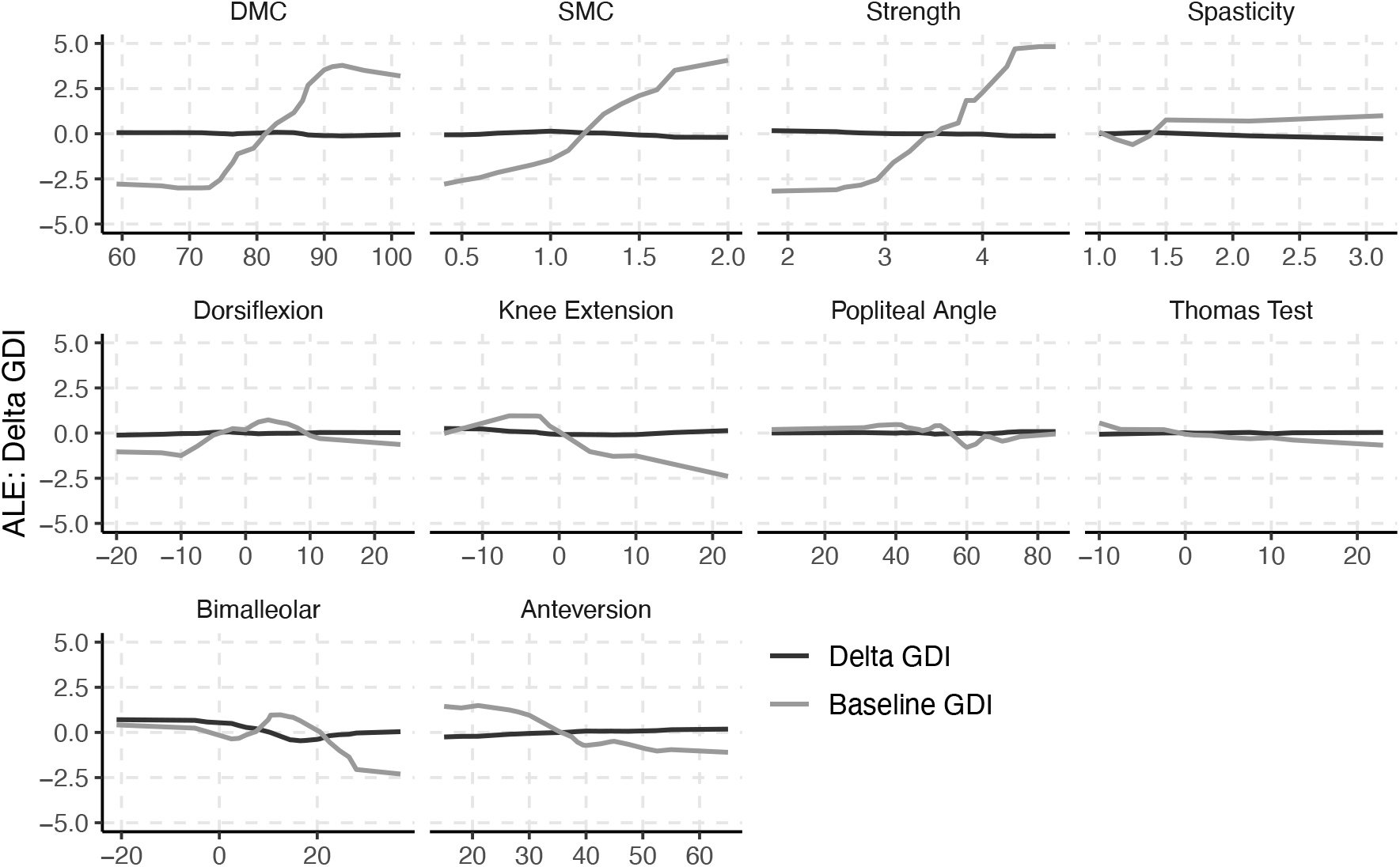
Accumulated local effects (ALE) of baseline neurologic and orthopedic impairments on GDI_pre_ and ΔGDI.

## 4 Discussion

This study showed that SEMLS has a small positive causal effect on change in GDI for children with bilateral CP. A ten-point change in GDI is generally considered a clinically significant improvement in walking function^57^. The observed change in GDI and total causal effect were far below this threshold. However, the cohorts who did not undergo SEMLS experienced a reduction in GDI between visits, resulting in a net effect of SEMLS around 1.68 GDI points. While average changes in GDI were modest, there was significant variation in outcome between participants, which could not be predicted by the model including baseline age, impairment level, or surgical history. We found that SEMLS produced an increase in GDI larger than five points for 35% of participants, but also a decrease of more than five points in 20% of the participants. Such heterogeneous responses to SEMLS have motivated our team’s investigations into patient-specific factors that can improve outcomes for children with CP. We ultimately want to be able to determine why an individual walks the way they do and anticipate their responses to treatment. We had previously hypothesized that motor control could be once such factor.

Our prior retrospective regression analyses demonstrated that DMC was *associated* with GDI after treatment across analyses at multiple clinical centers^5,6^. However, using a causal model to control for and evaluate the relative effects of various impairments indicated that motor control and other impairments had minimal *causal* effects on change in GDI. Importantly, DMC and other impairments had a strong effect on baseline GDI, indicating that these impairments do have a causal effect on a child’s gait pattern. However, these impairments had minimal effect on ΔGDI. In other words, a child who has greater DMC at baseline was likely to have a higher GDI than a child with lower DMC, but better motor control had minimal effect on expected changes in GDI. An important point in these analyses is that the overall causal effect of SEMLS was small, which contributes to the small observed effects of impairments on ΔGDI. Despite these small treatment effects, the wide heterogeneity in outcomes suggests that there are still causal factors contributing to treatment outcome that we are missing. These may include post-operative rehabilitation, surgeon skill, or other measures of neurologic impairment. Identifying patient-specific factors that can help us understand the causal pathways that impact gait and treatment outcomes continues to be an important area for future research.

Causal modeling provides a framework to evaluate the complex relationships between impairments and outcomes in CP. We created a DAG to identify the assumed relationships between SEMLS and GDI. The DAG used in this research could be expanded to include more detail about the assumed causal relationships between specific neurologic and orthopedic impairments or to evaluate other outcome measures. Similarly, our goal in this research was *not* to make outcome predictions for individual patients. Rather, we wanted to understand the impact of SEMLS and impairments on GDI. This led us towards more coarse modeling choices. As an example, we ignored details of surgical procedures and did not attempt to define the causal relationships between various neurologic and orthopedic impairments, although this is an area for future study.

The DAG we created for this research gave rise to the adjustment sets necessary to evaluate the impact of SEMLS and impairments on GDI. These adjustment sets can be used with any modeling method, including linear regression or other machine learning methods. We selected BART rather than linear regression or other models because we do not expect the impact of many impairments on gait to be linear. For example, we expect impairments like tibial torsion to reduce GDI scores with excessive internal or external rotation, producing a “u-shaped” response. Similarly, for some impairments like spasticity, there may be a threshold above or below which the impairment has an effect on gait. BART also provides a Bayesian framework that gives posterior distributions for each parameter.

A limitation in this research was that we did not recruit a prospective control group. Rather, we identified participants who were evaluated at multiple CGAs without any intervening surgical procedures. This cohort may also be subject to sample bias, but randomization is not feasible for this population. Since we were interested in evaluating DMC measured from EMG recordings, we also excluded children who underwent rectus femoris transfer, since the impact of moving the insertion of this muscle on recruitment and synergies remains unclear. Thus, this sample may not capture the impact of impairments that influence stiff-knee gait in children with CP.

### 4.1 Conclusions

The overall causal effect of SEMLS on change in GDI is modest. While motor control and strength do influence an individual’s gait pattern, their effect on expected changes in GDI after SEMLS are small. It is important to consider causal frameworks when analyzing observational data to avoid bias arising from confounding. Critically evaluating current CGA practices and integrating measures such as postoperative care, surgical details, or neuroimaging into treatment planning may enhance our ability to perform casual analyses aimed at understanding and improving movement for children with CP.

## Data Availability

Data produced in the present study are available upon reasonable request to the authors.

