## Supplementary Material for "Causal effects of motor control on gait kinematics after orthopedic surgery in cerebral palsy: a machine-learning approach"

#### 1 Directed Acyclic Graph

In this study we evaluate the impact of SEMLS (exposure) on ΔGDI (outcome).

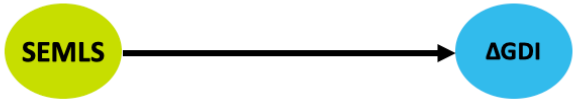


The decision to have a SEMLS does not directly cause a change in gait kinematics. Rather, ΔGDI is mediated by a change in impairments (ΔImp) caused by the surgery.

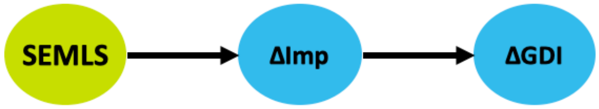


Baseline age, impairment level ($Imp_{pre}$), and $GDI_{pre}$ have a causal effect on these factors. Age also effects $Imp_{pre}$ and $GDI_{pre}$. Baseline impairment also has a causal effect on $GDI_{pre}$.

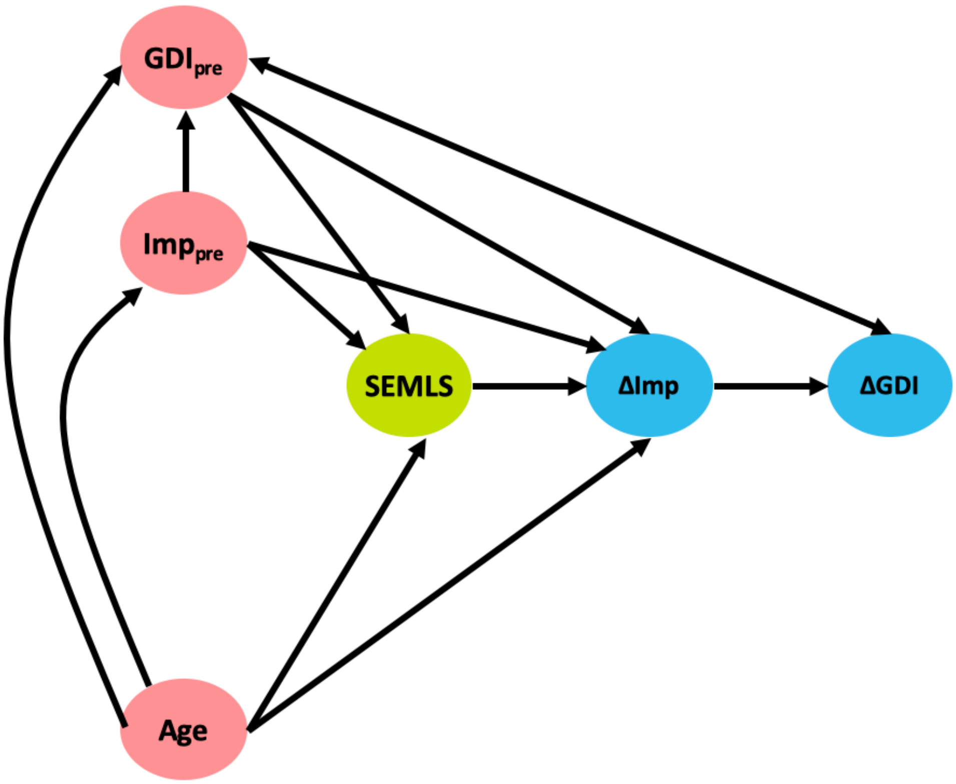

Prior surgical history (Hx) also impacts the decision to undergo SEMLS and baseline impairment levels. We also recognize that there are unmeasured factors related to an individual's impairment severity (Sev) that impact their history and baseline impairment.

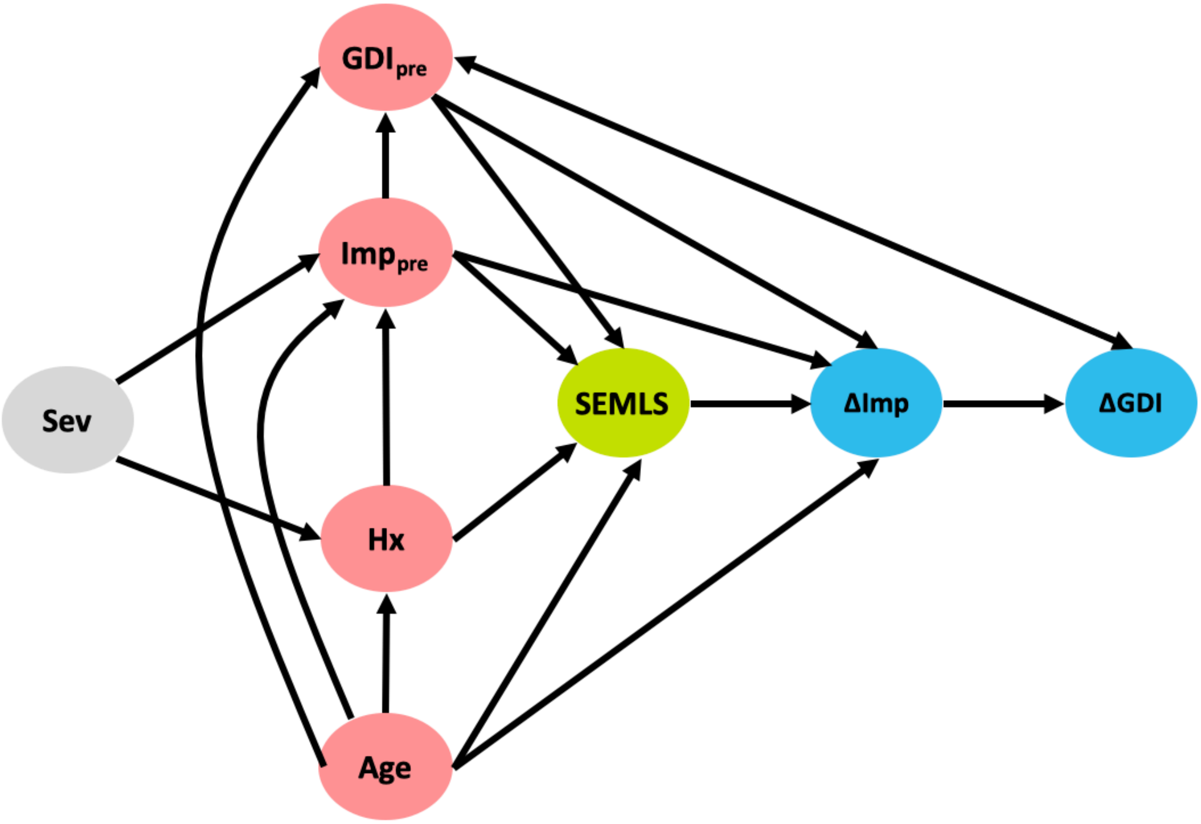

### 2 Missing Data

Using data collected as part of clinical care often means that a values are missing for a given visit. We used the R Multivariate Imputation by Chained Equations Package (MICE), which uses Fully Conditional Specification, a method where a separate model is developed for each variable to impute missing data.

**Table 1:** Number of participants missing each study variable

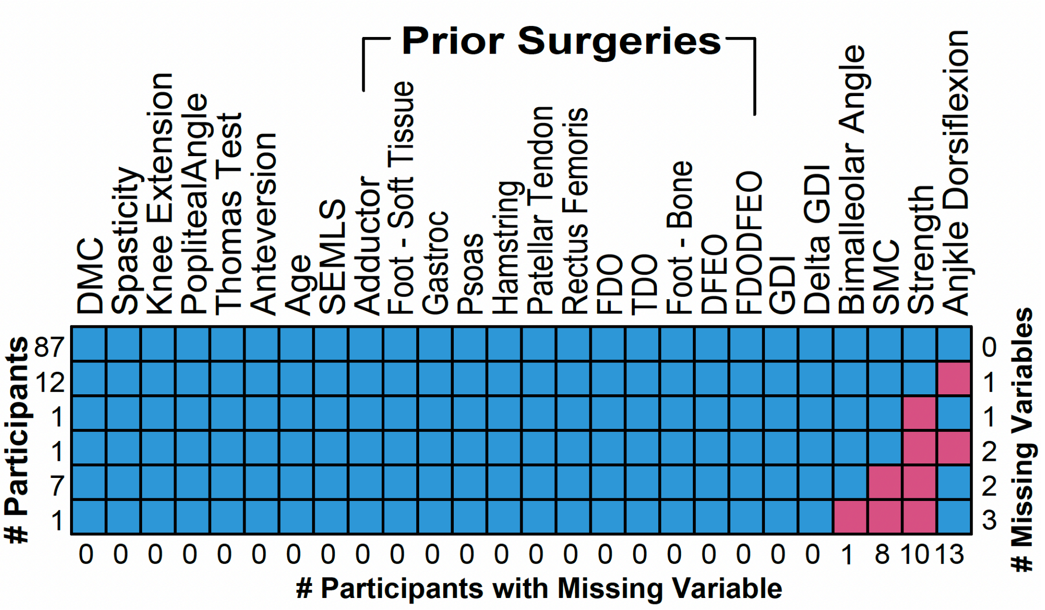


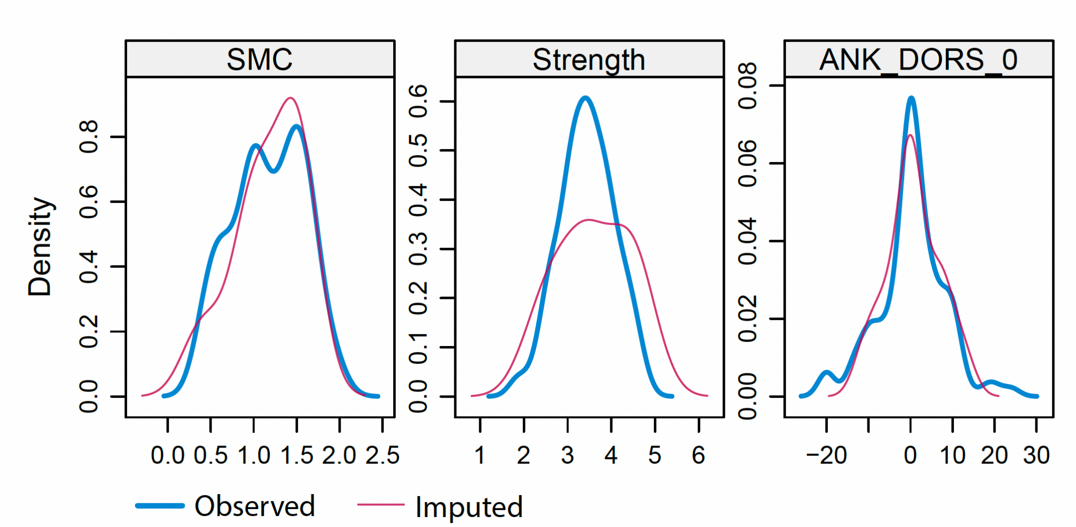


**Figure 1:** Density plots of study variables that had missing values. The blue line shows the observed distribution of each variable among participants with available data, and the red line shows the distribution of imputed missing data. SMC: Selective Motor Control.

### 3 Implied Conditional Independencies

We evaluated the implied conditional independencies specified from the assumed causal relationships defined in our DAG. All partial correlations were less than 0.20, supporting model plausibility.

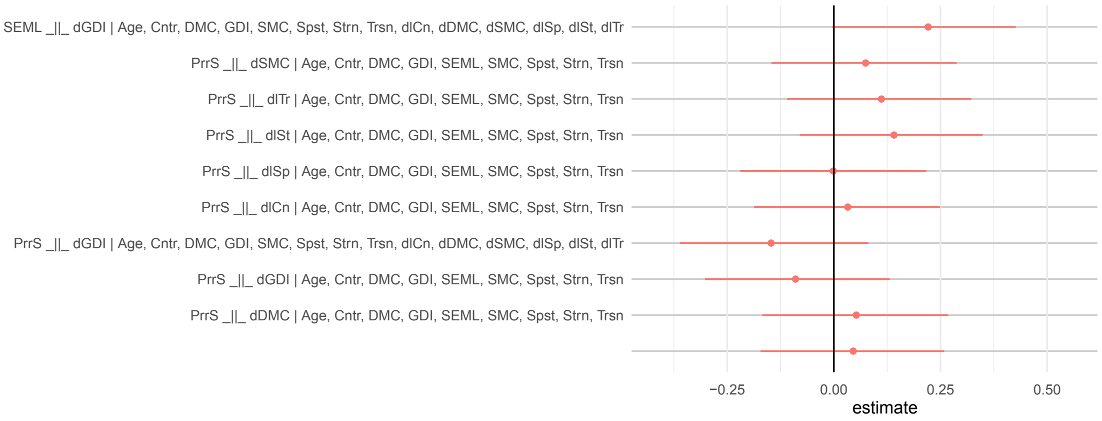
